# Low Dose Naltrexone Prescribing Practices for Children and Adolescents with Long COVID

**DOI:** 10.64898/2026.02.20.26346719

**Authors:** Cindy Villatoro, Alexandra B Yonts, Thomas Barter, Sindhu Mohandas, Laura A. Malone

## Abstract

**Background:** Pediatric long COVID is associated with substantial symptom burden, yet evidence-based pharmacologic treatments remain limited. Low-dose naltrexone (LDN) has been proposed as a potential symptomatic therapy, but data in pediatric populations is lacking.

**Methods:** We conducted a retrospective analysis of pediatric and young adult patients (≤25 years) with a clinical diagnosis of long COVID who were prescribed LDN between July 2020 and July 2025 at three multidisciplinary pediatric long COVID programs in the United States. Deidentified clinical data were extracted from medical records. Outcomes included symptom prevalence, dosing practices, treatment continuation or discontinuation, adverse effects, and available patient-reported quality-of-life measures (PedsQL™ and PROMIS®).

**Findings:** The study included 62 patients (mean age, 15.6 years [range, 8–23]; 53.2% male and 46.8% female). Fatigue was nearly universal (98.4%), followed by headaches (87.1%), brain fog (74.2%), dizziness/lightheadedness (67.7%), anxiety (66.1%), and post-exertional malaise (56.5%). LDN-treated patients demonstrated a higher prevalence of neurocognitive and autonomic symptoms, compared to general clinic cohorts. Most patients (71.0%) reported no adverse effects; the most common were vivid dreams (9.7%) and insomnia (9.7%). At follow-up, 66.1% of patients remained on LDN. Medication discontinuation was attributed to perceived lack of benefit (43.8%) or side effects (25.0%). Baseline quality-of-life measures at initiation showed marked impairment: PedsQL Physical Health (*M=*38.0, *SD*=20.9) and Multidimensional Fatigue (*M=*35.7, *SD*=15.8) scores were low. PROMIS scores indicated reduced physical functioning (*M*=36.8, *SD*=8.7) and cognitive functioning (*M*=40.8, *SD*=7.6), with elevated fatigue *(M*=68.0, *SD*=10.4) and pain interference *(M*=58.6, *SD*=8.2) relative to population norms. The study was not designed to assess efficacy.

**Interpretation:** LDN was primarily prescribed to patients with prominent fatigue, neurocognitive symptoms, and autonomic dysfunction, and was generally well tolerated. These findings provide descriptive evidence of real-world prescribing practices and support the need for clinical trials to systematically evaluate LDN’s efficacy in pediatric long COVID.

## INTRODUCTION

A subset of children and adolescents infected with SARS-CoV-2 experience recurrent symptoms following the acute phase of illness, a condition known as long COVID or post-acute sequelae of SARS-CoV-2 infection (PASC).^1^ Pediatric populations remain understudied, resulting in gaps in epidemiologic characterization, symptom profiling, and treatment guidance.^2^ Diagnosis is further complicated by fluctuating symptoms, limited pediatric-specific diagnostic criteria, and variability across developmental stages, making identification and timely management challenging for clinicians.^2–4^

Prevalence estimates for pediatric long COVID vary between 1% to 25%.^5–7^ Certain demographic groups, particularly adolescent females and Hispanic children, are disproportionately affected.^6,8^ Symptom presentations include fatigue, cognitive difficulties, headaches, dizziness, gastrointestinal symptoms, autonomic dysfunction, musculoskeletal pain, and sleep disturbances.^5,7,9^ These symptoms often persist for an extended time and are associated with disruptions in school participation, daily functioning, and overall quality of life.^5,8,10^

Currently, there are no Food and Drug Administration (FDA)-approved therapeutics for long COVID, as its underlying mechanisms remain under investigation.^11^ A symptomatic management approach is used in both adult and pediatric populations.^12,13^ Clinicians and families are exploring repurposed medications with potential anti-inflammatory or immunomodulatory effects, as some evidence suggests signs of immune dysregulation and chronic inflammation as potential mechanisms underlying long COVID in a subset of patients.^11^

Low-dose naltrexone (LDN), typically prescribed at 1–4.5 mg/day, has gained interest following early adult long COVID studies reporting improvements in fatigue, sleep disturbance, and post-exertional malaise, with good medication tolerance.^1,14^ Naltrexone is an opioid antagonist, and at higher doses (50-100mg), it is an FDA-approved medication for opioid and alcohol use disorder treatment to reduce cravings and prevent euphoric effects of alcohol and opioids. ^15^Naltrexone and its active metabolite, 6-beta-naltrexone, pharmacologically blocks the mu-opioid receptor, with weaker antagonistic action against the kappa and delta-opioid receptors.^15^ Recently, it has been used to treat binge eating and purging in adults and adolescents, thought to be effective by modulating neural reward system functioning.^16–18^

Although data on LDN for pediatric long COVID are limited, studies of other pediatric conditions suggest its potential safety and effectiveness. A pilot trial and retrospective reviews indicate favorable tolerability and symptom relief in some youth with Crohn’s disease and chronic pain.^19,20^ The findings highlight LDN’s suitability for pediatric use and its potential benefits for pain and fatigue in children with long COVID. The National Institutes of Health RECOVER–Treating Long COVID (RECOVER-TLC) program further reflects growing national interest in evaluating the safety and effectiveness of targeted therapies, including potential therapeutic candidates such as LDN for pediatric long COVID.^21^

To address the current evidence gap in pediatric populations, this retrospective observational study describes a cohort of patients younger than 25 years evaluated across three pediatric long COVID clinics in the United States who have previously or are currently trialing LDN for symptom management. This study characterizes demographics, presenting symptoms, dosing patterns, reported clinical responses, and side effects to inform future research and ongoing national initiatives evaluating LDN as a potential therapeutic strategy for pediatric long COVID.

## METHODS

### Study Design and Setting

We conducted a multicenter, retrospective case series of pediatric and young adult patients diagnosed with long COVID. Data were contributed by three pediatric long COVID programs in the United States (Kennedy Krieger Institute [KKI], Baltimore, MD; Children’s National Hospital [CNH], Washington, DC; and Children’s Hospital Los Angeles [CHLA], Los Angeles, CA). Each participating site obtained institutional review board approval to share deidentified data for secondary research, and a consent waiver was granted due to the use of routinely collected clinical information.

### Participants

The study included patients aged 25 years or younger who received clinical care for long COVID at one of the three sites between July 2020 and July 2025. Patients were eligible if they had a clinical diagnosis of long COVID based on provider evaluation and documentation of prior or current LDN prescribed for symptom management. Patients were excluded if their symptoms were unlikely to be related to SARS-CoV-2 infection or if they had insufficient medication records to confirm LDN initiation or treatment course.

### Data Sources and Measures

Eligible cases were identified through electronic medical records and specific registries. Study personnel gathered demographic and clinical data, including age at first evaluation, sex, race/ethnicity, date of COVID-19 infection, and pre-existing conditions. Long COVID symptoms were extracted from clinical notes and intake forms. LDN-related data included initiation date, starting dose, titration schedule, dose adjustments, reasons for discontinuation, and documented side effects.

### Clinical Safety and Tolerability

The primary outcome was treatment tolerability and clinical response. Adverse events and patient-reported reasons for discontinuing LDN were documented in provider notes and categorized as ineffective, side effects, symptom resolution, or unknown/not reported. This data was collected as part of routine care and reflects side effects reported to clinicians during clinical encounters related to maintenance or modification of LDN, rather than from standardized e-diaries for medication tracking. Some patients completed standardized patient-reported outcome measures as part of routine clinical care. KKI administered the Pediatric Quality of Life Inventory (PedsQL™) Generic Core and Fatigue Scales, and CNH collected Patient-Reported Outcomes Measurement Information System (PROMIS®) pediatric self-report and parent-proxy measures assessing physical functioning/mobility, cognitive functioning, fatigue, and pain interference.

The PedsQL™ Generic Core and Multidimensional Fatigue Scales assess physical, psychosocial, and fatigue-related functioning using child self-report (ages 5–18) and parent-proxy (ages 2–18) formats, both on a 5-point Likert scale; higher scores indicate better functioning and lower fatigue burden.^10,22,23^ Additional outcomes were measured using PROMIS® pediatric self-report and parent-proxy instruments (ages 8–17), which generate standardized T-scores (mean = 50, SD = 10).^24^ Score directionality varies by domain: higher values on symptom-focused scales (e.g., fatigue, pain interference) indicate greater symptom burden, whereas higher values on functioning scales (e.g., physical mobility) reflect better health status. ^24,25^

### Statistical Analysis

Descriptive methods were used to summarize all study variables. Demographic characteristics, comorbidities, symptom patterns, and LDN medication side effects were summarized using counts and percentages. Age and treatment duration were summarized using measures of central tendency. Patient-reported outcome scores from PedsQL™ and PROMIS® measures were also summarized descriptively to characterize overall functioning and symptom impact at PROMIS® self-report and parent-proxy measures. All analyses were performed using Microsoft Excel and SPSS (IBM Corporation).

## RESULTS

### Demographic and Clinical Characteristics

A total of 62 pediatric and young adult patients were included **(Table 1)**. The mean age was 15.6 years (SD 2.95; range 8–23 years), and most were adolescents. Slightly more than half were male (53.2%). The majority identified as White (71.0%), with smaller representation from other racial and ethnic groups. Approximately one-third of patients (32.3%) had no documented pre-existing conditions. The most common comorbidities were anxiety (24.2%), attention-deficit/hyperactivity disorder (ADHD) (22.6%), asthma (12.9%), and depression (11.3%).

**Table 1.** Baseline Demographic and Clinical Characteristics of the Cohort (N = 62). Demographic characteristics and pre-existing medical conditions reported by at least 2 participants are shown; most patients had no pre-existing medical conditions.

| Demographics | Count (%) |
| --- | --- |
| Sex |  |
| Male | 33 (53.2%) |
| Female | 29 (46.8%) |
| Age |  |
| 8-12 years | 10 (16.1%) |
| 13-17 years | 35 (56.5%) |
| 18+ years | 17 (27.4%) |
| Race/Ethnicity |  |
| White/Caucasian | 44 (71.0%) |
| Asian | 4 (6.5%) |
| More than one race | 4 (6.5%) |
| Black/African American | 2 (3.2%) |
| American Indian or Alaska Native | 1 (1.6%) |
| Other | 3 (4.8%) |
| Unknown/Not specified | 4 (6.5%) |
| Hispanic or Latino | 6 (9.7%) |
| **Pre-existing Conditions** |  |
| None | 20 (32.3%) |
| Anxiety | 15 (24.2%) |
| ADHD | 14 (22.6%) |
| Asthma | 8 (12.9%) |
| Depression | 7 (11.3%) |
| Seasonal allergies | 7 (11.3%) |
| Migraines | 4 (6.5%) |
| Concussions | 5 (8.1%) |
| OCD | 4 (6.5%) |
| Sensory Processing Disorder | 3 (4.8%) |
| Food and Medication Allergies | 3 (4.8%) |
| Ehlers-Danlos Syndrome | 2 (3.2%) |
| Constipation | 2 (3.2%) |
| Hypermobility | 2 (3.2%) |
| Generalized Anxiety Disorder | 2 (3.2%) |

### Long COVID Symptom Burden and Site-Specific Variation

Fatigue was reported by 98.4% of patients. Other common symptoms include headaches (87.1%), brain fog (74.2%), dizziness/lightheadedness (67.7%), anxiety (66.1%), concentration difficulties (61.3%), and post-exertional malaise (56.5%) **(Figure 1A)**. Over half of patients experienced nausea (54.8%), shortness of breath (50.0%), and heart racing/palpitations (50.0%), with 66.1% reporting 10 or more symptoms.

**Figure 1:**
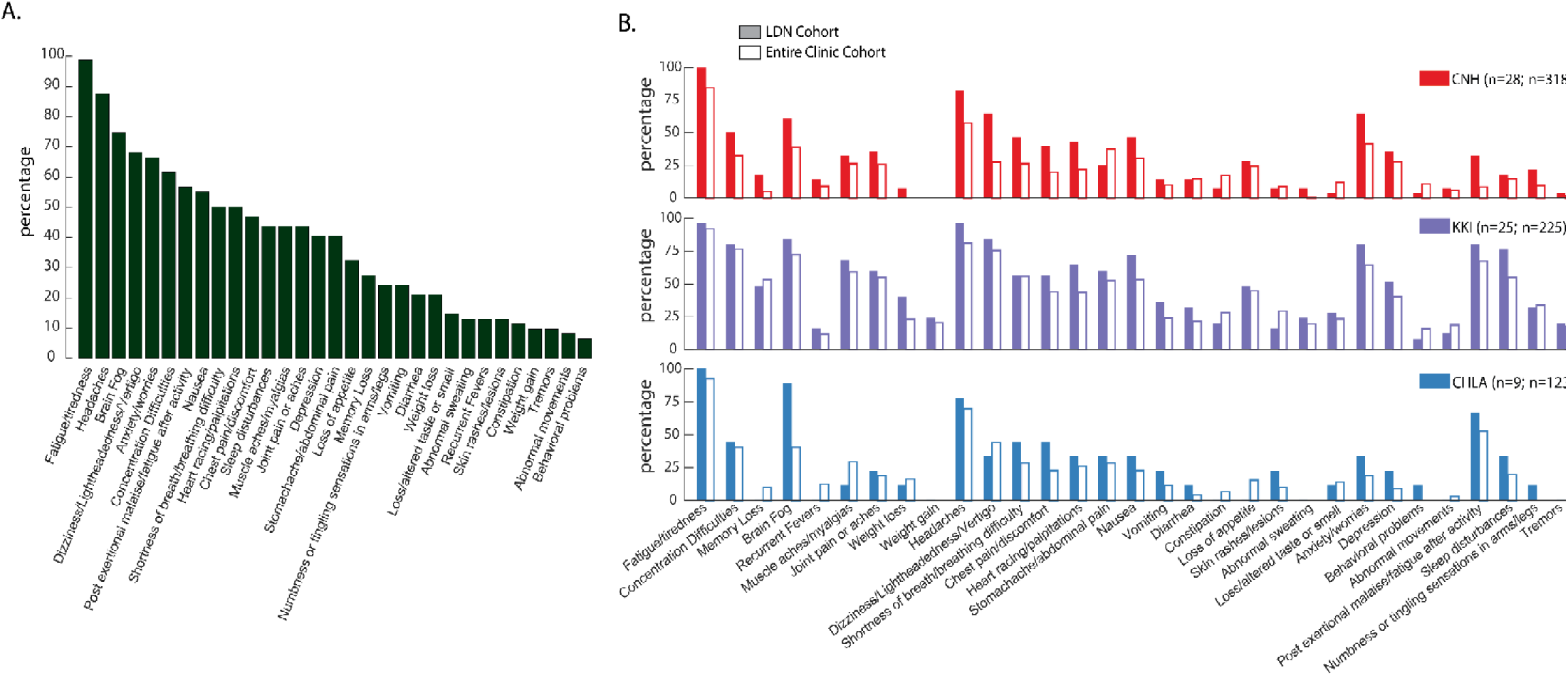
Distribution of long COVID Symptoms in the Full Cohort and by Clinical Site. **(A)** Distribution of reported long COVID symptoms in the full cohort (*N* = 62; CNH *n* = 28; KKI *n* = 25; CHLA *n* = 9), shown as the percentage of patients endorsing each symptom at clinical evaluation. **(B)** Symptom prevalence by clinical site. Colored bars represent symptom prevalence among patients prescribed low-dose naltrexone (LDN), and adjacent white bars represent symptom prevalence in the full clinic cohort at each site (CNH *N* = 318; KKI *N* = 225; CHLA *N* = 123), which includes patients with long COVID who were not prescribed LDN. **Notes:** In the full clinic cohort, data for weight gain and weight loss were not available at CNH. At CHLA, data were not available for weight gain, abnormal sweating, behavioral problems, numbness or tingling in the arms or legs, and tremors. Other symptoms without bars are reported as 0%.

Among patients prescribed LDN, symptom prevalence differed from their respective clinical cohorts across participating sites **(Figure 1B)**. Fatigue was nearly universal among LDN-treated patients (96–100%) compared to the full clinic populations (90–93%). Post-exertional malaise was also more common in LDN patients, particularly at CNH (32.1% vs. 8.5%). Neurocognitive symptoms, especially brain fog, were reported more frequently among LDN patients than in each clinic population (e.g., CNH: 60.7% vs. 38.7%; CHLA: 88.9% vs. 40.7%; KKI: 84.0% vs. 72.0%). Headaches were likewise more prevalent (e.g., 82.1% vs. 57.5% at CNH). Autonomic symptoms, especially dizziness/lightheadedness, were reported at higher rates among study patients at CNH (64.3% vs. 27.3%) and KKI (84.0% vs. 75.6%). Joint pain (22.2%–60.0%) and abdominal pain (25.0%–60.0%) showed the widest ranges. Overall, LDN was primarily prescribed to patients with prominent fatigue, post-exertional malaise, neurocognitive impairment, and autonomic symptoms.

### LDN Treatment Patterns

The mean time between symptom onset or initial SARS-CoV-2 infection and initiation of LDN was 21.59 months (SD 13.79), with a median of 20 months (range 0–54). Providers titrated doses gradually, with most patients reaching a daily dose of 4.5 mg. The most common maintenance dose was 4.5 mg/day (71.0%), followed by 1.5 mg (11.3%) and 3 mg (9.7%). Prescriptions for doses below 4.5 mg were attributed to maximum patient tolerance, effectiveness at lower doses without the need to increase, or ongoing titration at the time of data collection.

During data collection, 41 patients (66.1%) remained on LDN **(Figure 2).** Sixteen patients (25.8%) discontinued treatment, mainly due to perceived lack of benefit (43.75%), medication side effects (25.0%), and symptom resolution (18.8%); 12.5% had unknown reasons or unreported data. One patient had not yet initiated the medication. Some patients reported no symptom improvement with LDN but experienced worsening symptoms upon discontinuation, leading to therapy re-initiation.

**Figure 2.**
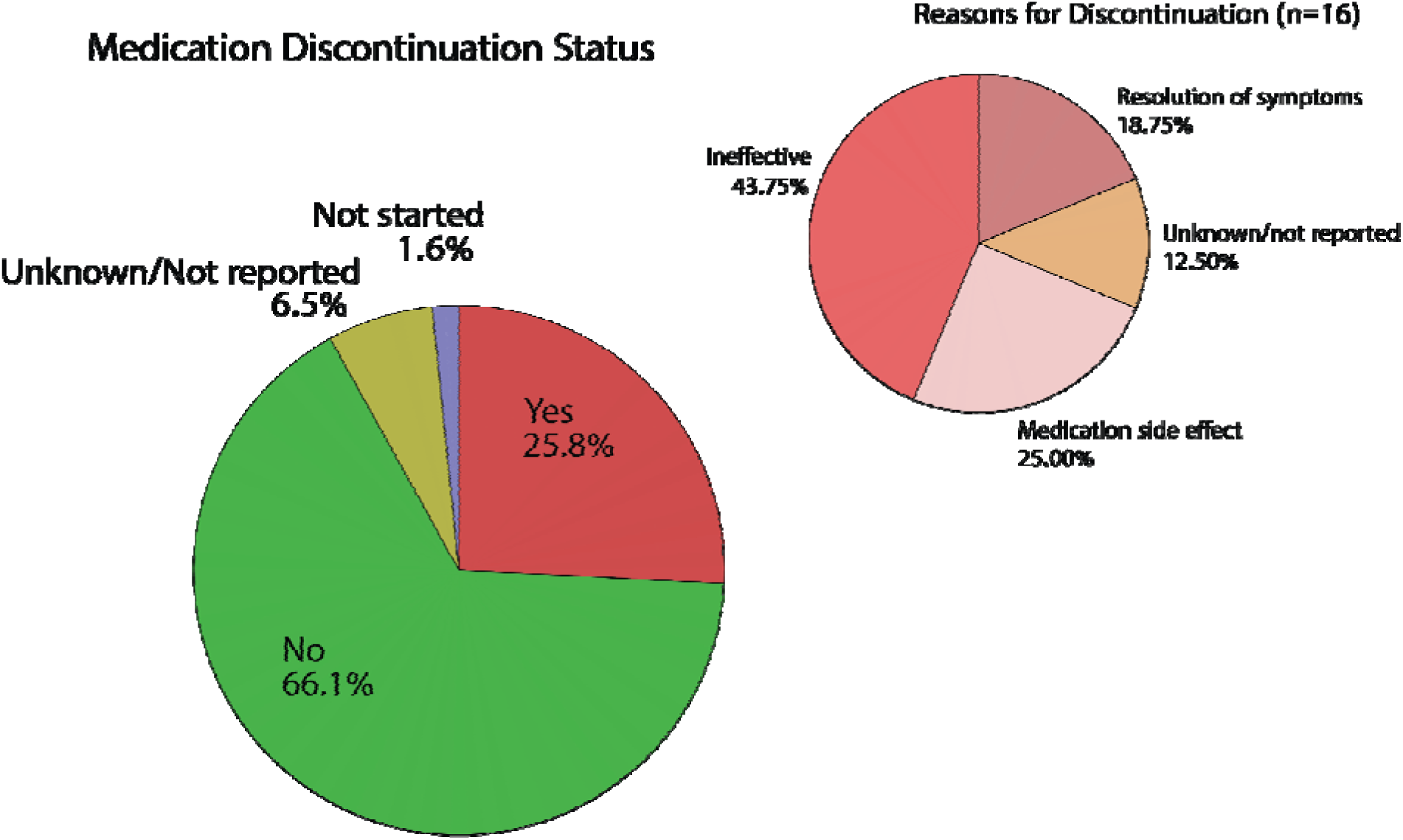
LDN Discontinuation Status and Reasons for Discontinuation. Two pie charts summarizing LDN treatment status in the cohort. The left chart depicts the proportion of patients who continued, discontinued, or had unknown initiation/discontinuation status. The upper right chart illustrates the proportion of documented reasons among those who discontinued treatment.

### Safety and Tolerability

LDN was generally well tolerated, with 71.0% of patients experiencing no adverse effects. The most common reported side effects were vivid dreams/nightmares (9.7%), difficulty sleeping/insomnia (9.7%), fatigue (6.5%), headaches/migraines (6.5%), and dizziness/lightheadedness (3.2%) **(Table 2)**. Notably, some side effects may overlap with long COVID symptoms, suggesting reported effects could be attributed to long COVID rather than LDN. However, patients’ perceived medication side effects are presented for completeness.

**Table 2:** Patient-Reported Medication Side Effects from LDN.

| **Medication Side Effects** | **Count (%)** |
| --- | --- |
| None | 44 (71.0%) |
| Vivid dreams/Nightmares | 6 (9.7%) |
| Difficulty sleeping/Insomnia | 6 (9.7%) |
| Fatigue | 4 (6.5%) |
| Headaches/Migraines | 4 (6.5%) |
| Dizziness/Lightheadedness | 2 (3.2%) |
| Syncopal episodes | 2 (3.2%) |
| Worsening pain | 1 (1.6%) |
| Excessive eye blinking | 1 (1.6%) |
| Exacerbation of GI symptoms | 1 (1.6%) |
| Arms and legs tingling | 1 (1.6%) |
| Muscle twitching | 1 (1.6%) |
| Brain fog | 1 (1.6%) |
| Behavioral challenges | 1 (1.6%) |
| Nausea | 1 (1.6%) |
| Unknown/Not reported | 1 (1.6%) |

### Quality of Life Outcomes

Standardized patient-reported outcome measures were available for a subset of patients at two participating sites around the time of LDN initiation **(Figure 3)**. PedsQL™ Generic Core scores indicated reductions in health-related quality of life, affecting both physical health (Physical Health Summary Score of *M* = 38.0 (*SD* = 20.9) and psychosocial health (Psychosocial Summary Score of *M* = 51.7 (*SD* = 12.5). Fatigue-related functioning reflected low levels of Cognitive Fatigue (*M* = 42.97, *SD* = 19.4) and Overall Multidimensional Fatigue (*M* = 35.72, *SD* = 15.77).

**Figure 3.**
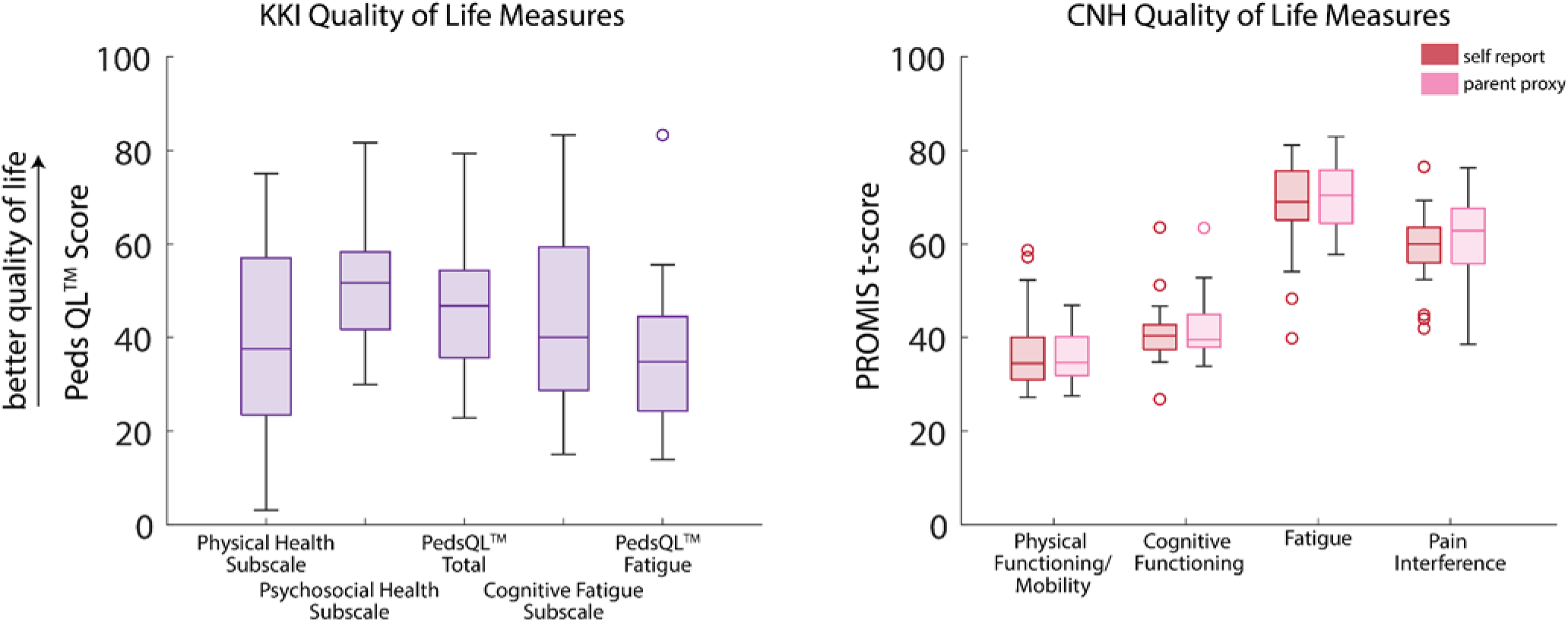
Quality-of-Life Scores at Visits Closest to LDN Initiation. Box-and-whisker plots summarizing patient-reported quality-of-life measures from the visit closest to LDN initiation. **Left:** Pediatric Quality of Life Inventory (PedsQL) Generic Core and Fatigue Scale scores from the KKI cohort. High values indicate a better quality of life. **Right:** Patient-Reported Outcomes Measurement Information System (PROMIS) self-report and parent-proxy T-scores for physical functioning, cognitive functioning, fatigue, and pain interference from the CNH cohort. For the Physical Functioning/Mobility and Cognitive Functioning PROMIS measures, higher scores indicate better functioning or improved quality of life. For the Fatigue and Pain Interference PROMIS measures, higher scores indicate worse impairment/symptoms.

PROMIS® T-scores from the CNH cohort demonstrated a similar pattern. Scores were similar between the self-report and parent proxy PROMIS® scores (Physical functioning/mobility: r=0.79, p<0.001, Cognitive functioning: r=0.70, p=0.003, Fatigue: r=0.72, p<0.001, Pain Interference: r=0.57, p=0.01). Physical functioning/mobility T-scores were well below the population norm (self-report *M* = 36.75, *SD* = 8.74; parent-proxy *M* = 35.51, *SD* = 5.35), and cognitive functioning scores were likewise reduced (self-report *M* = 40.79, *SD* = 7.56; parent-proxy *M* = 41.68, *SD* = 7.54). In contrast, fatigue scores were elevated, indicating greater symptom burden (self-report *M* = 68.04, *SD* = 10.43; parent-proxy *M* = 69.68, *SD* = 7.46). Pain interference scores were also elevated relative to expected norms (self-report *M* = 58.55, *SD* = 8.21; parent-proxy *M* = 61.12, *SD* = 8.94), indicating meaningful impacts on daily functioning. These findings demonstrate baseline impairments across physical, cognitive, fatigue, and pain domains at the time LDN treatment was initiated; however, the effect of LDN on quality of life was not systematically studied here.

## DISCUSSION

Using retrospective data from three pediatric long COVID programs in the US, this study characterizes the clinical features and treatment context of children, adolescents, and young adults prescribed LDN. While this cohort is unique to multidisciplinary specialty care, the experience with LDN reported in this study is likely generalizable to the broader pediatric long COVID population. The demographic composition, including age, sex, and race, mirrors that of large US observational studies. However, a lower percentage of patients receiving LDN identified as Hispanic/Latino, compared to observational cohorts (9.7% vs 15-22%).^2,26^ The over-representation of White, non-Hispanic patients in specialty care settings has been reported in long COVID and other Infection-Associated Chronic Conditions (IACC) clinical settings.^27,28^ This disparity likely reflects systemic barriers to healthcare access and the geographic restriction of specialized clinics.^27,28^

Over half (68%) of the cohort had at least one pre-existing medical condition; common diagnoses included anxiety, ADHD, asthma, depression, and seasonal allergies, which are also reported in pediatric long COVID cohorts, though at lower frequencies.^26^ The predominant symptoms in patients receiving LDN were consistent with general reporting of pediatric long COVID, with high rates of fatigue, cognitive dysfunction, dizziness/lightheadedness, anxiety, and post-exertional malaise. Differences in symptomatic indication and burden across sites (Figure 1) may reflect variations in prescribing practices, referral patterns, or patient populations, and may also indicate heterogeneity in clinical expression, as pediatric long COVID likely comprises subgroups with differing symptom profiles and severity. This symptom pattern is consistent with reported indications for LDN in adult long COVID populations and other neurocognitive conditions. ^14,16–18^ Patients receiving LDN tended to be more severely affected by their symptoms at baseline, per patient-reported outcome measures (PROMIS®), compared to other pediatric long COVID cohorts,^26^ particularly in the domains of pain, fatigue, and physical functioning. On average, patients initiated LDN therapy almost 2 years after SARS-CoV2-infection/symptom onset, likely reflecting delays in diagnosis and care access, and inadequate response to non-pharmacologic interventions, such as pacing, lifestyle modifications, and cognitive or physical therapy.^29,30^

The mechanism by which LDN may alleviate symptoms in long COVID and other conditions, such as fibromyalgia or myalgic encephalomyelitis/chronic fatigue syndrome, remains incompletely understood. Proposed hypotheses include action via anti-inflammatory or immunomodulatory effects. Different doses of naltrexone in animal models have opposing effects.^31^ For instance, low doses of naltrexone inhibit tumor growth, while high-dose naltrexone can accelerate tumor growth and somatic cell development.^32^ One hypothesis is that immunomodulatory effects of LDN occurs via intermittent or partial blockade of the opioid growth factor receptor (originally called □ [zeta] receptor), displacing opioid growth factor (met-enkephalin).^33^ This intermittent blockade leads to altered T-cell, natural killer cell, and macrophage activity and adjustments in cytokine production such as IL-1, IL-6, TNF-alpha and Interferon-gamma.^34^ The *duration* of blockade is particularly important (as opposed to the dose) for the immunomodulatory effects along the opioid growth factor (OGF)-OGF receptor (OGFr) axis’s role^33^ in cell proliferation for development and homeostasis.^34^ Other proposed mechanisms of immunomodulatory effects, such as toll-like receptor 4 antagonism and glial cell modulation are seen at much higher, supratherapeutic concentrations of naltrexone in preclinical studies.^35–38^

Optimal dosing and the effects of different dosing ranges on the mechanisms of LDN action in humans remain major questions. Gradual dose escalation is a consistent aspect across published studies,^39–41^ although the starting doses (0.1mg - 1.5mg) and target maintenance dose (3.0mg - 6.0mg) remain variable. Although gradual dose escalation is commonly reported, the dose increase per time point varies. In this study, most patients initiated therapy at 1.5mg and achieved a maximum dose of 4.5mg/day without severe adverse effects. Additionally, the primary reason for discontinuing LDN was the perceived lack of clinical improvement rather than adverse effects.

It remains unclear whether the adverse effects of LDN are dose-related, as this study did not systematically assess dose–response relationships (maximum 4.5mg/day). Further research is needed to determine whether higher doses provide greater efficacy without increased side effects. A notable challenge in evaluating tolerability was distinguishing medication side effects from fluctuating long COVID symptoms, particularly fatigue, headaches, and lightheadedness, which overlap with known LDN side effects.^42^ However, new onsets of insomnia and vivid dreams or nightmares after LDN initiation were more clearly attributable to the medication and are well-documented side effects.^42^

Another challenge with LDN is its limited availability, as it can only be obtained through compounding pharmacies. Commercially, it is available only as a 50 mg tablet for opioid use disorder treatment. LDN formulations are prepared by specialized compounding pharmacies in personalized doses ranging from 0.5 mg to 6 mg, most commonly as immediate-release capsules and tablets. Anecdotally, most patients in this study used the capsule form of LDN. LDN’s proposed mechanism of action entails brief opioid receptor blockade and an endorphin rebound effect, making immediate-release formulations preferable to achieve the effect, and extended-release preparations should be avoided. Fillers used in capsule formulations can also impact the rate of naltrexone absorption; therefore, consistent preparation is crucial to avoid variability, especially for clinical trials. It is recommended that LDN formulations be sourced from experienced compounding pharmacies.^31,41,43^

In this multi-site retrospective cohort study, we are unable to determine the efficacy of LDN directly on pediatric long COVID symptoms; however, anecdotally, many patients report improvement in symptoms after initiating therapy. This study was conducted as part of routine clinical care, with follow-up periods irregularly scheduled based on patient symptoms, functioning, and availability. Factors such as reinfection and concurrent therapies could also confound interpretations of efficacy, and this data was not collected for this study.

Despite these limitations, this study demonstrates the safety and tolerability of LDN in young patients with long COVID and describes common prescribing practices amongst pediatric long COVID experts. Across sites, there was consistency in prescribing LDN to more severely affected patients who reported fatigue, headaches, and neurocognitive symptoms such as brain fog and difficulty concentrating, suggesting that the efficacy might be best in this population.

Additional placebo-controlled clinical trials are needed, as planned by the RECOVER-TLC initiative, to directly address the many remaining questions regarding the efficacy of LDN for specific symptomatology or phenotypic presentations of pediatric long COVID, and to more accurately describe age-specific side-effect profiles.

## Data Availability

Data produced in the present study are available upon reasonable request to the corresponding author.

## Data sharing statement

Deidentified individual participant data and related study documents will not be made publicly available. Requests for data access may be directed to the corresponding author, Laura Malone, MD, PhD.

## REFERENCES

1. Isman A, Nyquist A, Strecker B, Harinath G, Lee V, Zhang X, et al. Low-dose naltrexone and NAD+ for the treatment of patients with persistent fatigue symptoms after COVID-19. Brain Behav Immun - Health. 2024 Mar;36:100733.

2. Gross RS, Thaweethai T, Kleinman LC, Snowden JN, Rosenzweig EB, Milner JD, et al. Characterizing Long COVID in Children and Adolescents. JAMA. 2024 Oct 8;332(14):1174.

3. Liu VY, Godfrey M, Dunn M, Fowler R, Guthrie L, Dredge D, et al. Diagnostic challenges of long COVID in children: a survey of pediatric health care providers’ preferences and practices. Front Pediatr. 2024 Dec 23;12:1484941.

4. Gross RS, Thaweethai T, Salisbury AL, Kleinman LC, Mohandas S, Rhee KE, et al. Characterizing Long COVID Symptoms During Early Childhood. JAMA Pediatr. 2025 Jul 1;179(7):781.

5. Henning E, Musci R, Johnson SB, Villatoro C, Malone LA. Pediatric long COVID: relationships with premorbid history of anxiety or depression and health-related quality of life. J Pediatr Psychol. 2025 Oct 1;50(10):919–26.

6. Vahratian A, Ph.D., M.P.H., Adjaye-Gbewonyo D, Ph.D., M.P.H., et al. Products - Data Briefs - Number 479 - September 2023 [Internet]. 2025 [cited 2026 Jan 17]. Available from: https://www.cdc.gov/nchs/products/databriefs/db479.htm

7. Kamel D, Vu MH, Bender J, Warburton D, Wood JC, Mohandas S. Exploring long COVID in pediatric patients: clinical insights from a long COVID clinic. Front Pediatr. 2025 Oct 15;13:1640747.

8. Basaca DG, Jugănaru I, Belei O, Nicoară DM, Asproniu R, Stoicescu ER, et al. Long COVID in Children and Adolescents: Mechanisms, Symptoms, and Long-Term Impact on Health—A Comprehensive Review. J Clin Med. 2025 Jan 9;14(2):378.

9. Toepfner N, Brinkmann F, Augustin S, Stojanov S, Behrends U. Long COVID in pediatrics—epidemiology, diagnosis, and management. Eur J Pediatr. 2024 Jan 27;183(4):1543–53.

10. Chen EY, Morrow AK, Malone LA. Exploring the Influence of Preexisting Conditions and Infection Factors on Pediatric Long COVID Symptoms and Quality of Life. Am J Phys Med Rehabil. 2024 Jul;103(7):567.

11. Peluso MJ, Deeks SG. Mechanisms of long COVID and the path toward therapeutics. Cell. 2024 Oct;187(20):5500–29.

12. Malone LA, Morrow A, Chen Y, Curtis D, de Ferranti SD, Desai M, et al. Multi-disciplinary collaborative consensus guidance statement on the assessment and treatment of postacute sequelae of SARS-CoV-2 infection (PASC) in children and adolescents. PM&R. 2022;14(10):1241–69.

13. Cheng AL, Herman E, Abramoff B, Anderson JR, Azola A, Baratta JM, et al. Multidisciplinary collaborative guidance on the assessment and treatment of patients with Long COVIDC: A compendium statement. PM&R. 2025 Jun;17(6):684–708.

14. Bonilla H, Tian L, Marconi VC, Shafer R, McComsey GA, Miglis M, et al. Low-dose naltrexone use for the management of post-acute sequelae of COVID-19. Int Immunopharmacol. 2023 Nov;124:110966.

15. Singh D, Saadabadi A. Naltrexone. In: StatPearls [Internet]. Treasure Island (FL): StatPearls Publishing; 2025 [cited 2026 Jan 17]. Available from: http://www.ncbi.nlm.nih.gov/books/NBK534811/

16. Stancil SL, Abdel-Rahman S, Wagner J. Developmental Considerations for the Use of Naltrexone in Children and Adolescents. J Pediatr Pharmacol Ther. 2021;26(7):675–95.

17. Stancil SL, Voss M, Nolte W, Tumberger J, Adelman W, Abdel-Rahman S. Effects of genotype and food on naltrexone exposure in adolescents. Clin Transl Sci. 2022 Nov;15(11):2732–43.

18. Stancil SL, Yeh HW, Brucks MG, Bruce AS, Voss M, Abdel-Rahman S, et al. Potential biomarker of brain response to opioid antagonism in adolescents with eating disorders: a pilot study. Front Psychiatry. 2023 Jul 10;14:1161032.

19. Smith JP, Field D, Bingaman SI, Evans R, Mauger DT. Safety and Tolerability of Low-dose Naltrexone Therapy in Children With Moderate to Severe Crohn’s Disease: A Pilot Study. J Clin Gastroenterol. 2013 Apr;47(4):339–45.

20. Theriault C, Oyelola O, Zempsky WT. The Efficacy Of Low-Dose Naltrexone In Pediatric Chronic Pain: A Retrospective Analysis. J Pain. 2023 Apr 1;24(4):84–5.

21. RECOVER-TLC invites public comments on upcoming clinical trials | RECOVER COVID Initiative [Internet]. [cited 2026 Jan 17]. Available from: https://recovercovid.org/news/recover-tlc-invites-public-comments-upcoming-clinical-trials

22. Varni JW, Limbers CA, Burwinkle TM. Impaired health-related quality of life in children and adolescents with chronic conditions: a comparative analysis of 10 disease clusters and 33 disease categories/severities utilizing the PedsQL^TM^ 4.0 Generic Core Scales. Health Qual Life Outcomes. 2007 Dec;5(1):43.

23. Varni JW, Limbers CA, Bryant WP, Wilson DP. The PedsQL^TM^ Multidimensional Fatigue Scale in type 1 diabetes: feasibility, reliability, and validity. Pediatr Diabetes. 2009 Aug;10(5):321–8.

24. Luijten MAJ, Haverman L, Van Litsenburg RRL, Roorda LD, Grootenhuis MA, Terwee CB. Advances in measuring pediatric overall health: the PROMIS® Pediatric Global Health scale (PGH-7). Eur J Pediatr. 2022 May;181(5):2117–25.

25. DeWalt DA, Gross HE, Gipson DS, Selewski DT, DeWitt EM, Dampier CD, et al. PROMIS® pediatric self-report scales distinguish subgroups of children within and across six common pediatric chronic health conditions. Qual Life Res. 2015 Sep;24(9):2195–208.

26. Montealegre Sanchez GA, Arrigoni LE, Yonts AB, Rubenstein KB, Bost JE, Wolff MT, et al. Pediatric SARS-CoV-2 long term outcomes study (PECOS): cross sectional analysis at baseline. Pediatr Res. 2025 Aug;98(2):541–50.

27. Smyth N, Ridge D, Kingstone T, Gopal DP, Alwan NA, Wright A, et al. People from ethnic minorities seeking help for long COVID: a qualitative study. Br J Gen Pract. 2024 Dec;74(749):e814–22.

28. Shi J, Lu R, Tian Y, Wu F, Geng X, Zhai S, et al. Prevalence of and factors associated with long COVID among US adults: a nationwide survey. BMC Public Health. 2025 May 13;25(1):1758.

29. Au L, Capotescu C, Eyal G, Finestone G. Long covid and medical gaslighting: Dismissal, delayed diagnosis, and deferred treatment. SSM - Qual Res Health. 2022 Dec 1;2:100167.

30. Gu L, Yue J, Lin J, Liu Z, Huang J an. Challenges in diagnosis and treatment of long COVID. Front Med. 2025 Aug 21;12:1641411.

31. Toljan K, Vrooman B. Low-Dose Naltrexone (LDN)—Review of Therapeutic Utilization. Med Sci. 2018 Sep 21;6(4):82.

32. Li Z, You Y, Griffin N, Feng J, Shan F. Low-dose naltrexone (LDN): A promising treatment in immune-related diseases and cancer therapy. Int Immunopharmacol. 2018 Aug;61:178–84.

33. McLaughlin PJ, Zagon IS. Duration of opioid receptor blockade determines biotherapeutic response. Biochem Pharmacol. 2015 Oct;97(3):236–46.

34. Hankins GR, Harris RT. The Opioid Growth Factor in Growth Regulation and Immune Responses in Cancer. In: Kerr PL, Sirbu C, Gregg JM, editors. Endogenous Opioids [Internet]. Cham: Springer International Publishing; 2024 [cited 2026 Jan 21]. p. 45–85. (Advances in Neurobiology; vol. 35). Available from: https://link.springer.com/10.1007/978-3-031-45493-6_4

35. Cabanas H, Muraki K, Staines D, Marshall-Gradisnik S. Naltrexone Restores Impaired Transient Receptor Potential Melastatin 3 Ion Channel Function in Natural Killer Cells From Myalgic Encephalomyelitis/Chronic Fatigue Syndrome Patients. Front Immunol. 2019 Oct 31;10:2545.

36. Hutchinson MR, Zhang Y, Brown K, Coats BD, Shridhar M, Sholar PW, et al. Non-stereoselective reversal of neuropathic pain by naloxone and naltrexone: involvement of toll-like receptor 4 (TLR4). Eur J Neurosci. 2008 Jul;28(1):20–9.

37. Cant R, Dalgleish AG, Allen RL. Naltrexone Inhibits IL-6 and TNFα Production in Human Immune Cell Subsets following Stimulation with Ligands for Intracellular Toll-Like Receptors. Front Immunol. 2017 Jul 11;8:809.

38. Wang X, Zhang Y, Peng Y, Hutchinson MR, Rice KC, Yin H, et al. Pharmacological characterization of the opioid inactive isomers (+)-naltrexone and (+)-naloxone as antagonists of toll-like receptor 4. Br J Pharmacol. 2016 Mar;173(5):856–69.

39. Marcus N, Robbins L, Araki A, Gracely E, Theoharides T. Effective Doses of Low-Dose Naltrexone for Chronic Pain – An Observational Study. J Pain Res. 2024 Mar;Volume 17:1273–84.

40. O’Kelly B, Vidal L, McHugh T, Woo J, Avramovic G, Lambert JS. Safety and efficacy of low dose naltrexone in a long covid cohort; an interventional pre-post study. Brain Behav Immun - Health. 2022 Oct;24:100485.

41. Younger J, Parkitny L, McLain D. The use of low-dose naltrexone (LDN) as a novel anti-inflammatory treatment for chronic pain. Clin Rheumatol. 2014 Apr;33(4):451–9.

42. Aalto H, Paul S, McEwen V. Real-World Effectiveness and Tolerability of Low Dose Naltrexone to Treat Chronic Pain: A Retrospective Cohort Study of One Pain Physician’s Practice. J Pain Res. 2025 Dec;Volume 18:6637–49.

43. LDN Research Trust - The Low Dose Naltrexone Charity [Internet]. [cited 2026 Jan 17]. Available from: https://ldnresearchtrust.org/

